# Individual prediction of trauma-focused psychotherapy response in youth with posttraumatic stress disorder using resting-state functional connectivity

**DOI:** 10.1101/2021.01.11.21249596

**Authors:** Paul Zhutovsky, Jasper B. Zantvoord, Judith B.M. Ensink, Rosanne Op den Kelder, Ramon J.L. Lindauer, Guido A. van Wingen

**Affiliations:** Amsterdam UMC, University of Amsterdam, Department of Psychiatry, Amsterdam Neuroscience, Amsterdam, The Netherlands; Amsterdam UMC, University of Amsterdam, Department of Child and Adolescent Psychiatry, Amsterdam Neuroscience, Amsterdam, The Netherlands; De Bascule, Academic Centre for Child and Adolescent Psychiatry, Amsterdam, The Netherlands; Research Institute of Child Development and Education, University of Amsterdam, Amsterdam, The Netherlands

## Abstract

Randomized controlled trials have shown efficacy of trauma-focused psychotherapies in youth with posttraumatic stress disorder (PTSD). However, response varies considerably among individuals. Currently, no biomarkers are available to assist clinicians in identifying youth who are most likely to benefit from treatment. In this study, we investigated whether resting-state functional magnetic resonance imaging (rs-fMRI) could distinguish between responders and non-responders on the group- and individual patient level.

Pre-treatment rs-fMRI was recorded in 40 youth (ages 8-17 years) with (partial) PTSD before trauma-focused psychotherapy. Change in symptom severity from pre- to post-treatment was assessed using the Clinician-Administered PTSD scale for Children and Adolescents to divide participants into responders (≥30% symptom reduction) and non-responders. Functional networks were identified using meta-independent component analysis. Group-differences within- and between-network connectivity between responders and non-responders were tested using permutation testing. Individual predictions were made using multivariate, cross-validated support vector machine classification.

A network centered on the bilateral superior temporal gyrus predicted treatment response for individual patients with 76.17% accuracy (p_FWE_ = 0.018, 87.14% sensitivity, 65.20% specificity, area-under-receiver-operator-curve of 0.82). Functional connectivity between the frontoparietal and sensorimotor network was significantly stronger in non-responders (p_FWE_ = 0.012) on the group-level. Within-network connectivity was not significantly different between groups.

This study provides proof-of-concept evidence for the feasibility to predict trauma-focused psychotherapy response in youth with PTSD at an individual-level. Future studies are required to test if larger cohorts could increase accuracy and to test further generalizability of the prediction models.

## Introduction

Posttraumatic stress disorder (PTSD) is a common mental health disorder that develops in approximately 16% of youth exposed to traumatic events, which may include domestic violence, sexual abuse and accidents [1]. Youth with PTSD are troubled by frequent re-experiencing of the traumatic event, persistent avoidance, hyperarousal and negative alterations in cognition and mood [2]. These symptoms can interfere with social functioning and school performance and have a negative effect on the quality of life of the affected youth and their families [3]. Moreover, they are a crucial factor in shaping the vulnerability to depression and suicidality later in life [4].

Multiple randomized controlled trials (RCTs) have demonstrated the efficacy of trauma-focused psychotherapies in youth with PTSD [5]. However, response varies considerably among individuals, with 30-50% of youth who do not benefit sufficiently from treatment [5,6], leading to persistent PTSD symptoms, comorbidity and longer treatment trajectories. Although different pre-treatment clinical and demographic factors have been associated with trauma-focused psychotherapy outcome, none has shown to consistently distinguish between responders and non-responders or reliably predict treatment response [7]. This underlines the need for the identification of reliable (bio)markers of treatment response which could assist clinicians to optimize treatment allocation to improve clinical outcome.

Previous studies utilizing (functional) magnetic resonance imaging ((f)MRI) have shown that PTSD is characterized by alterations in brain regions which are key nodes in multiple large-scale brain networks [8]. The current neurocircuitry model of PTSD in adults suggests hyperactivity and increased connectivity within the salience network (SN) involving the amygdala and anterior insula [9]. The SN is responsible for detecting and orienting to salient stimuli [10]. Furthermore, PTSD patients have shown reduced activity and connectivity within the default mode network (DMN) [11], a network associated with internally focused thought as well as autobiographical memory and includes the ventromedial prefrontal cortex and the posterior cingulate cortex [10]. A third frontoparietal network (FPN) or central executive network centered around the dorsolateral prefrontal cortex and lateral parietal cortex is associated with goal-directed behavior, decision making and working memory [10]. Previous studies in PTSD have shown reduced connectivity within the FPN [12]. Together these findings suggest that adult PTSD is characterized by distinct dysfunctional patterns in large-scale networks which may contribute to abnormal saliency towards trauma-related material, working memory problems and difficulties in processing trauma related material.

Importantly, results from studies examining large-scale network connectivity in youth with PTSD have not always corresponded with results obtained in adults [13]. Patriat, et al. ^14^, for instance, found that pediatric PTSD is characterized by increased connectivity within the DMN, contrasting the finding of decreased connectivity within the DMN in adults. This could be related to the fact that large-scale brain networks undergo considerable reorganization throughout childhood and adolescence [13]. In general, developmental change in large-scale brain organization is characterized by stronger within network connectivity and more efficient between-network connectivity, with a trend towards segregation (decrease in connectivity strength) between regions in close proximity and integration (increase in connectivity strength) between anatomically distant regions [15]. More specifically, age-related reductions in strength between the DMN, SN and FPN resulting in more segregated functioning of these networks [15]. These developmental processes provide a potential explanation for the contrasting findings between youth and adults with PTSD [13] and emphasize the need for studies on large-scale brain networks specifically performed in youth with PTSD.

Interestingly, relatively few studies have investigated the relationship between treatment-response and large-scale brain network connectivity. In adults with PTSD pre-treatment difference between trauma-focused psychotherapy responder and non-responder groups have been observed in activity and functional connectivity both during resting-state and task-based fMRI [16-20]. Findings from these studies suggest that activity and connectivity within regions and networks involved in working memory as well as emotional processing and modulation differed between responders and non-responders [16,17]. A study in adolescent girls with PTSD has reported greater pre-treatment bilateral amygdala activation in response to fear vs neutral facial expressions in treatment responders relative to non-responders and group differences in large-scale brain network connectivity during emotion processing [21]. These studies provide initial evidence for group-differences in pre-treatment brain activity and connectivity between treatment responders and non-responders.

These studies used univariate analysis focused on finding average group-differences. However, such an approach does not use new data to validate the observed finding and it remains unclear how such group-differences may be informative for the individual patients [22]. This is necessary to allow clinicians to inform patients and to assist in clinical decision making. Multivariate supervised machine learning analysis can be used to make predictions for individual patients instead. Several studies have utilized machine learning methods and rs-fMRI to predict treatment-response in adults with PTSD, with accuracies ranging between 71-90% [17,20,23]. However, to the best of our knowledge, there are no studies available that have investigated the utility of rs-fMRI to predict treatment-response for individual patients in youth with PTSD. Therefore, we collected pre-treatment rs-fMRI data of 40 youth with PTSD/partial-PTSD (age 8-17) to predict treatment response on the group- and individual-level.

## Materials and Methods

### Participants

Our initial sample consisted of 61 participants (39 female) diagnosed with PTSD or partial PTSD. Participants entered trauma-focused psychotherapy as part of a RCT comparing trauma-focused cognitive behavioral therapy (TF-CBT) and eye movement desensitization and reprocessing (EMDR) [6]. Of these, 50 completed treatment as well as pre- and post-treatment assessment (see flow diagram in Figure S1). After data quality control 40 participants (26 female) were included in the final analysis. All participants were Dutch speaking, and 8-17 years old. Participants were recruited between June 2011 and September 2018 at the outpatient child psycho-trauma center of the department of child and adolescent psychiatry, de Bascule in Amsterdam, The Netherlands. Youth were referred by child welfare services, physicians or general practitioners. Diagnoses for PTSD or partial PTSD were established clinically by an experienced child and adolescent psychiatrist or psychologists according to the DSM-IV-TR criteria using joint child and caregiver reports on individual symptoms on the Clinician-Administered PTSD Scale for Children and Adolescents (CAPS-CA) semi-structured interview [24] and the caregiver reports from the PTSD scale of the Anxiety Disorders Interview Schedule – Parent Version (ADIS-P) [25]. A symptom was established as present, if either child or caregiver reported its presence. Partial PTSD was defined as either fulfilling two of the three PTSD symptom clusters or having one symptom present in each of the three symptom clusters [26]. Furthermore, participants were required to have a CAPS-CA total score indicating at least mild PTSD symptom severity (>20 points). Exclusion criteria were: acute suicidality, IQ<70, pregnancy, neurological disorders or serious medical illnesses or meeting the criteria of the following diagnosis: psychotic disorders, substance-use disorder or pervasive developmental disorder. If participants were taking psychotropic or central nervous-active medication, medication was required to be stable for at least three weeks before and during trauma-focused psychotherapy. In our sample one participant was taking sertraline and two methylphenidate. In accordance with procedures approved by the Institutional Review Board of the Amsterdam University Medical Center and the declaration of Helsinki, written informed consent was obtained from all parents or legal guardians. Written informed consent from youth aged 12 years and older and assent from youth aged 11 and younger, was also obtained from the youth themselves. All participants received a monetary incentive for participation (€5 for each assessments).

### Trauma-focused psychotherapy

Participants were randomly assigned to eight weekly protocolized sessions of either TF-CBT or EMDR. The data reported here were obtained as part of a larger study on the long-term efficacy of TF-CBT and EMDR. This study is not yet unblinded as the long-term outcome is not yet available. But as these treatments share core aspects, such as in-vivo exposure to the traumatic memory and cognitive restructuring, and both are effective for youth with PTSD, we combined these treatments for the current analysis. Results using the same approach have been reported previously [27,28]. Treatment was delivered by experienced trauma therapists who were trained in TF-CBT and EMDR before study initiation. Supervision by TF-CBT and EMDR experts was provided throughout the study. Treatment protocols, training and supervision of therapists, as well as treatment fidelity have been described in detail previously [27].

Trained psychologists administered the CAPS-CA and the PTSD scale of the ADIS-P to measure PTSD symptoms before and after treatment. Caregiver reports on the ADIS-P were used to complement child reports and clinical observation. The Dutch Revised Child Anxiety and Depression Scale (RCADS(-P)) questionnaires was administered to assess depressive and anxiety symptoms [29]. Symptom change was calculated by subtracting the pre-treatment from the post-treatment CAPS-CA total score. We used ≥30% reduction of CAPS-CA total score as response criterion for clinically meaningful improvement [28].

The distribution of baseline clinical, trauma and demographic characteristics across responders and non-responders was examined using X^*2*^-tests, independent sample *t*-tests or Mann-Whitney tests as appropriate. Paired sample *t*-test were used to examine pre- to post-treatment symptom change. Statistical analyses were performed using SPSS version 26 (SPSS Inc., Chicago IL, USA).

### Imaging data acquisition

High-resolution T1 and rs-fMRI data were acquired using a 3T Philips Achieva scanner (Philips Healthcare, Best, The Netherlands) equipped with a SENSE eight-channel receiver head coil. For each participant, a T1-weighted structural MRI image was acquired with the following parameters: TE: 3.527 ms, TR: 9 ms, slice thickness: 1 mm, 170 slices, flip angle: 8° and image matrix 256 x 256 that covert the entire brain. 200 blood oxygen level dependent rs-fMRI scans were acquired with a repetition time of 2.3s and a voxel size of 2.3×2.3×3mm^3^. For rs-fMRI, participants were instructed to remain still with their eyes closed.

### Imaging data preprocessing

All (f)MRI data preprocessing was performed utilizing a singularity image container running fMRIPrep (v1.5.3, https://fmriprep.org/en/1.5.3/).

### Structural data preprocessing

Structural MR images were corrected for intensity non-uniformity and brain-extracted using the ANTs toolbox (v2.2.0, https://stnava.github.io/ANTs). Brain tissue segmentation of cerebrospinal fluid (CSF), white-matter (WM), and gray-matter (GM) was performed on the brain-extracted T1w images using FSL FAST (v5.0.9). Volume-based spatial normalization to MNI space (MNI152NLin6Asym) was performed through nonlinear symmetric normalization with ANTs.

### Functional data preprocessing

Preprocessing of rs-fMRI data followed the standard procedure implemented in fMRIPrep involving generation of a reference volume, co-registration to the T1w scan, motion correction (before any spatiotemporal filtering) and normalization to MNI space in one step using a combination of all spatial transformations (see Supplementary Materials for details). Normalizations and co-registrations were assessed visually and four PTSD patients were excluded due to poor normalization quality. We excluded five additional participants with high spikes of motion identified from visual inspection of plots of the realignment parameters (volume-to-volume changes >2mm). The remaining participants did not differ in overall motion levels according to their framewise displacement[30] (see Table 1). Data were spatially smoothed with an isotropic, Gaussian kernel of 6mm full-width-at-half-maximum. To further address motion contamination, we applied ICA-AROMA [31] (in MNI space) to remove additional motion sources from the data. Data was then resampled to 4mm^3^ to speed-up further computational steps. We addressed further structured noise present in the data by regressing out average WM and CSF signals using masks calculated in T1w space, transformed to rs-fMRI space. We combined this regression step with highpass filtering by a discrete cosine set with 128s cut-off. To avoid reintroducing already removed nuisance signal into the data by applying a sequential pipeline, both WM/CSF and cosine regressors were denoised with the previously identified ICA-AROMA regressors [32]. As a final step the rs-fMRI data were grand-mean scaled with a factor of 10000.

**Table 1.** Subject characteristics.

| **Table 1** Subject characteristics. | **Responders (n= 21)** | **Non-responders (n= 19)** | **p-value<sup>a</sup>** |
| --- | --- | --- | --- |
|  | **≥30% CAPS-CA** | **<30% CAPS-CA** |  |
| ***Sociodemographic characteristics*** |  |  |  |
| Female (%) | 57.1 | 73.7 | .273 |
| Age (years; mean, SD) | 12.5 (2.64) | 12.7 (3.25) | .820 |
| West European Ethnicity (%) | 52.4 | 42.1 | .413 |
| Current educational level (%) |  |  | .557 |
| Elementary school | 52.4 | 42.1 |  |
| Middle/High school lower level | 9.5 | 5.3 |  |
| Middle/High school middle level | 28.6 | 26.3 |  |
| Middle/High school higher level | 9.5 | 15.8 |  |
| Vocational school | 0 | 10.5 |  |
| Household Income (€; %) |  |  | .622 |
| <25000 | 28.6 | 26.3 |  |
| 25000-35000 | 19.0 | 5.3 |  |
| >35000 | 23.8 | 15.8 |  |
| Weight (kg; mean, SD) | 51.3 (12.67) | 50.7 (8.46) | .875 |
| Current psychotropic medication (%) | 9.5 | 5.3 | .609 |
| Smoking (%) | 9.5 | 5.3 | .702 |
| Alcohol >1 consumption/day (%) | 0 | 0 | N/A |
| ***Imaging Data*** |  |  |  |
| Framewise displacement (mean, SD) | 0.21 (0.11) | 0.20 (0.12) | .820 |
| ***Trauma characteristics*** |  |  |  |
| Index trauma (%) |  |  | .971 |
| Sexual abuse | 28.6 | 36.8 |  |
| Domestic violence | 14.3 | 10.5 |  |
| Community violence | 23.8 | 26.3 |  |
| Accidents/Medical | 14.3 | 10.5 |  |
| Other | 19.0 | 15.8 |  |
| Repeated trauma exposure (%) | 61.9 | 52.6 | .554 |
| Age at index trauma (years; mean, SD) | 10.0 (3.43) | 9.9 (4.42) | .824 |
| Time since index trauma (years; mean, SD) | 2.7 (2.00) | 2.9 (3.03) | .773 |
| ***Clinical characteristics*** |  |  |  |
| CAPS-CA study entry (mean, SD)<sup>b</sup> |  |  |  |
| Total | 55.5 (23.95) | 56.8 (23.09) | .856 |
| Re-experiencing | 16.7 (10.04) | 18.9 (10.92) | .532 |
| Avoidance | 22.8 (9.62) | 20.1 (10.54) | .422 |
| Hyperarousal | 16.8 (9.49) | 18.8 (8.48) | .515 |
| Full PTSD diagnosis (%) | 85.7 | 78.9 | .574 |
| RCADS study entry (mean, SD)<sup>b</sup> |  |  |  |
| MDD | 11.7 (6.11) | 12.5 (6.29) | .729 |
| GAD | 8.4 (4.56) | 5.6 (3.48) | .089 |
| OCD | 7.3 (3.84) | 6.2 (2.59) | .407 |
| PD | 9.2 (6.65) | 7.4 (5.81) | .469 |
| SAD | 7.4 (3.91) | 4.3 (4.12) | **.048** |
| SP | 13.3 (7.28) | 10.8 (6.00) | .339 |

Abbreviations: CAPS-CA, Clinician-Administered PTSD Scale for Children and Adolescents; RCADS, Revised Child Anxiety and Depression Scale; MDD, major depressive disorder; GAD, general anxiety disorder; OCD, obsessive compulsive disorder; PD, panic disorder; SAD, separation anxiety disorder; SP, social phobia; SE, standard error.
<sup>a</sup> $p$ -values <0.05 shown in bold. Independent samples t-test for continuous and $\chi^2$ tests for categorical variables.
<sup>b</sup> Ranges: CAPS-CA total, 0-139; RCADS MDD, 0-30; RCADS GAD, 0-18; RCADS OCD, 0-18; RCADS PD, 0-27; RCADS SAD, 0-21; RCADS SP, 0-27.

### Identification of intrinsic connectivity networks

To identify a set of robust intrinsic connectivity networks (ICNs) we employed a meta independent component analysis (ICA) [33] utilizing FSL MELODIC (v3.15) [34]. To ensure that the identification of ICNs was independent from their use in the analysis, which may introduce a positive bias [35], we included rs-fMRI data of 17 trauma-exposed controls (TEC) who did not differ in age, gender, or motion from the included patients (see Supplementary Materials for further details on the TEC and the meta-ICA). The number of components was fixed to 70 as it has been successful in the identification of treatment-related PTSD biomarkers for veterans in our previous study [17]. To identify ICNs, we employed a semi-automatic approach [36] which led to the inclusion of 48 ICNs (see Supplementary Materials). Both ICNs and excluded components are shown in Figure S2 and S3, respectively.

To reconstruct individual-level representations of the group-level ICNs and their time-courses we applied group-information guided ICA (GIG-ICA) to the preprocessed data of the PTSD patients [37]. GIG-ICA computes a spatially constrained individual-level ICA which takes the group-maps into account and estimates individual ICNs which are maximally spatially correlated with the corresponding group-map. This procedure is repeated for each group ICN and each participant, generating a set of individual-level ICN representations and their corresponding time-courses. GIG-ICA has been shown to outperform conventional reconstruction methods like dual regression in identifying reliable biomarkers for psychiatric disorders and to produce spatially independent components for the individual [37,38]. GIG-ICA was applied utilizing MATLAB code (R2018b, The Mathworks, Natick, MA) distributed with the GroupICA toolbox (v4.0b, https://trendscenter.org/software/gift).

To investigate between-ICN connectivity we applied the FSLnets toolbox (v0.6.3, https://fsl.fmrib.ox.ac.uk/fsl/fslwiki/FSLNets) to the individual-level ICN time-courses estimated via GIG-ICA. We estimated full- and partial-correlation matrices between all identified ICNs and converted all correlation coefficients to z-scores for further analyses. Only the unique part of the correlation matrices was considered in the analyses.

### Group-level analyses

We tested for group-differences across ICNs (within-ICN connectivity) between responders and non-responders using permutation testing implemented in PALM (a117, https://fsl.fmrib.ox.ac.uk/fsl/fslwiki/PALM). We included demeaned age, gender and pre-treatment CAPS-CA total scores as covariates-of-no-interest into a general linear model (GLM). Familywise error (FWE) correction of p-values across the whole-brain, 48 ICNs and two-sided tests of the threshold-free-cluster-enhancement (TFCE) statistic [39] was performed using synchronized permutations (n=10000) of the maximum statistic via PALM.

The same procedure, involving permutation testing (n=10000), and the same covariates-of-no-interest was utilized to investigate group-difference in between-ICN connectivity across responders and non-responders. The FWE-correction of p-values of the t-statistic was performed across all connections, two-sided contrasts and the full- and partial correlation matrices utilizing the maximum statistic. Alpha was set to 0.05 in both analyses.

### Individual-level analysis

To investigate whether within- or between-ICN connectivity could be utilized to predict treatment-response for the individual patient, we applied multivariate, cross-validated linear-kernel support vector classifiers (SVM) [40] to our data. For that we considered every ICN and their connectivity profiles (full- and partial-correlation matrices) separately, and divided our data into 5-folds (each fold containing 20% of the data) ensuring (approximate) balance of responders and non-responders per fold. Data of 4-folds was used as training set for rescaling all features to −1 to 1 range and fitting the SVM. The fifth fold served as the test set and we calculated balanced accuracy (average between sensitivity and specificity), area-under-the-receiver-operator-curve (AUC), sensitivity (of identifying responders), specificity (of identifying non-responders) and negative/positive predictive value (NPV/PPV) as performance measures for the trained SVM classifier. We repeated the procedure five times, each time retraining the classifier and utilizing a different fold as the test set. Finally, we repeated the division of the data across the five folds 50 times and repeated the entire analysis, yielding a 50-times-repeated-5-fold cross-validation procedure. In the end, we averaged the performance measures across the 250 test set evaluations, providing a set of measures estimating the generalizability of our classifier to new data.

To assess statistically whether the estimated average accuracies provided better-than-chance performance and to correct for the number of classifications performed, we used synchronized permutation tests (n=2000, see Supplementary Materials). Alpha was set to 0.05.

We also assessed which features were important for the classification by calculating p-values for each weight of the SVM using a novel permutation-based procedure [41] (see Supplementary Materials). The p-values were computed after the classifier was applied to the entire data set and are intended for visualization purposes only.

All individual-level analyses were implemented in the Python programming language (v3.8.2) utilizing the scikit-learn machine learning toolbox (v0.22.1).

## Results

### Demographic and clinical characteristics

A summary of participant characteristics is shown in Table 1. Treatment responders and non-responders did not differ in demographic, trauma and clinical characteristics at baseline apart from separation anxiety symptoms which were (marginally significantly) higher in responders (p=0.048). Based on joint child (CAPS-CA) and caregiver (ADIS-P) reports 82.5% of all participants met the full DSM-IV diagnostic criteria for PTSD at baseline, the remaining 17.5% met criteria for partial PTSD. The average baseline CAPS-CA score was 56.13 (SD=23.25), which is indicative of moderately severe PTSD. The most common index trauma was sexual abuse, followed by community violence, accidents and domestic violence. 57.5% of participants were exposed to multiple-event trauma. Average age at trauma exposure was M=9.95 years, SD=3.89 (range 2-16) and average time since trauma was M=2.82 years, SD=2.52 (range 0-10).

### Changes in psychopathology

Treatment completers and non-completers did not differ in baseline sociodemographic, trauma or clinical characteristics. Across the completer sample, we found significant reductions in CAPS-CA total score (*t*(39)=5.65, p<0.001, Cohen’s effect size (d)=0.89), re-experiencing (*t*(39)=4.39, p<0.001, d=0.71), avoidance (*t*(39)=4.10, p<0.001, d=0.68) and hyperarousal clusters (*t*(39)=2.935, p=0.006, d=0.55. Twenty-one fulfilled the criterion for treatment response (≥30% PTSD symptom reduction on CAPS-CA), and nineteen were non-responders.

### Resting-state fMRI

#### Group-level analyses

##### Within-network analyses

There were no group-differences surviving FWE-correction between responders and non-responders for any of the 48 ICNs. Because the number of investigated components was large, requiring stringent correction for multiple comparisons, we also provide the results of the analyses when FWE-correction was only applied for each network separately (see Figure S4). Within-network connectivity of two ICNs (left FPN and a visual ICN) was increased in responders over non-responders in this exploratory analysis.

##### Between-network analyses

Between-network analyses showed a significantly larger Pearson correlation between the (predominantly) left FPN and a sensorimotor network in non-responders over responders (p_FWE_=0.012, Figure 1).

**Figure 1:**
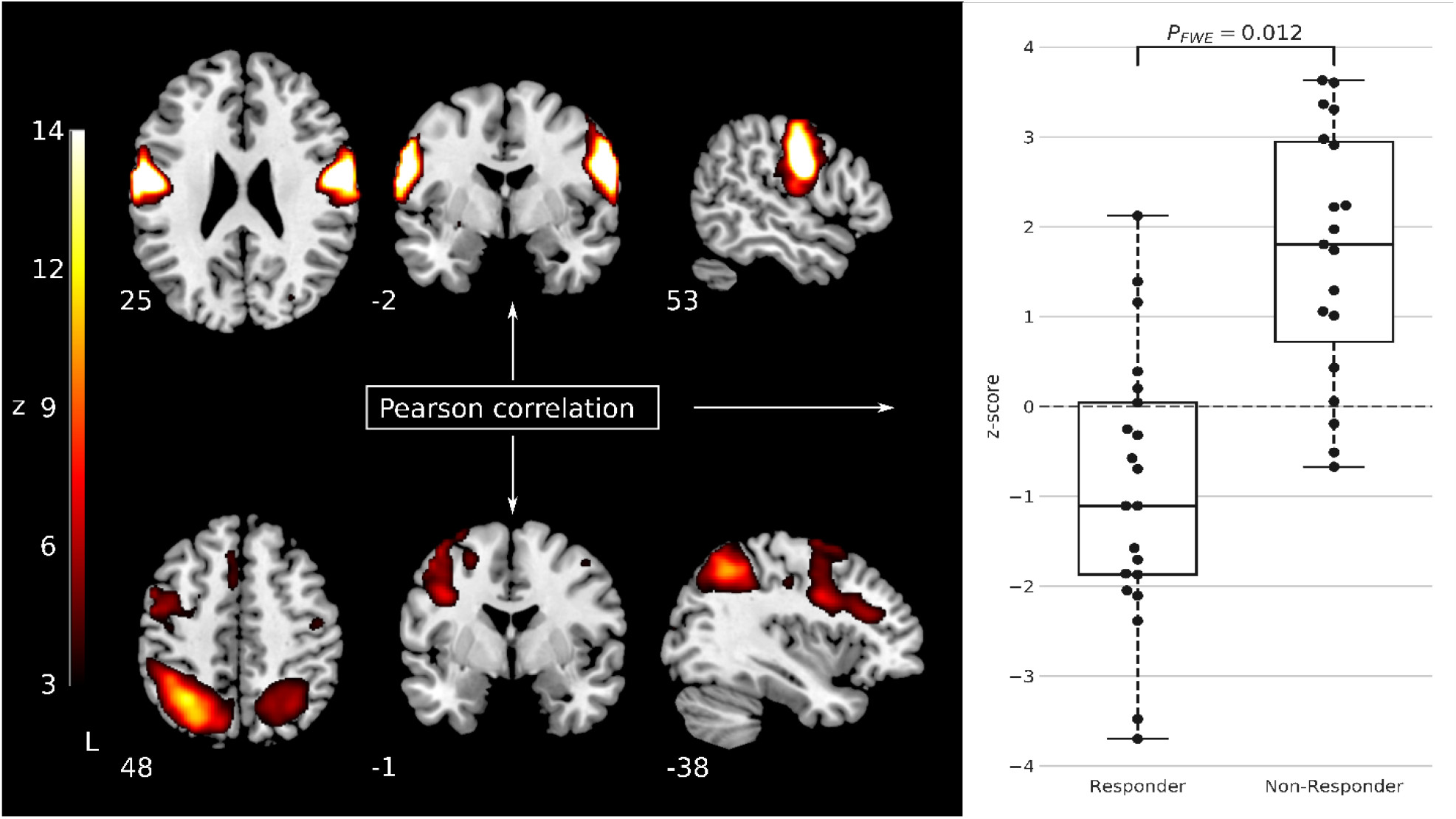
Stronger Fisher r-to-z transformed Pearson correlation between a sensorimotor network and the (predominately) left frontoparietal network was observed for non-responders over responders. Boxplots shows median and interquartile range of the distribution of responders/non-responders. The dots show the individual z-transformed correlation values of the individual patients.

### Individual-level analyses

SVMs trained on data from an ICN centered on the bilateral superior temporal gyrus (STG) provided an average cross-validated accuracy of 76.17% (SD=12.58%, p_FWE_=0.018, Figure 2(A) and Figure 3). The network achieved an AUC of 0.82 (SD=0.16), with a sensitivity of 87.14% (SD=16.56%) and a specificity of 65.20% (SD=21.44%). The PPV/NPV was 0.75/0.85 (SD=0.14/0.19). No other network showed classification accuracies exceeding chance-level when FWE-correction was applied. However, if no correction for the number of tests was performed, three more networks showed better-than-chance performance in the classification (Figure S5). P-values corresponding to the voxel-weights of the SVM classifier when trained on data of all patients of the STG ICN can be seen in Figure 2 (B), showing a diffuse whole-brain pattern required to successfully perform the classification.

**Figure 2:**
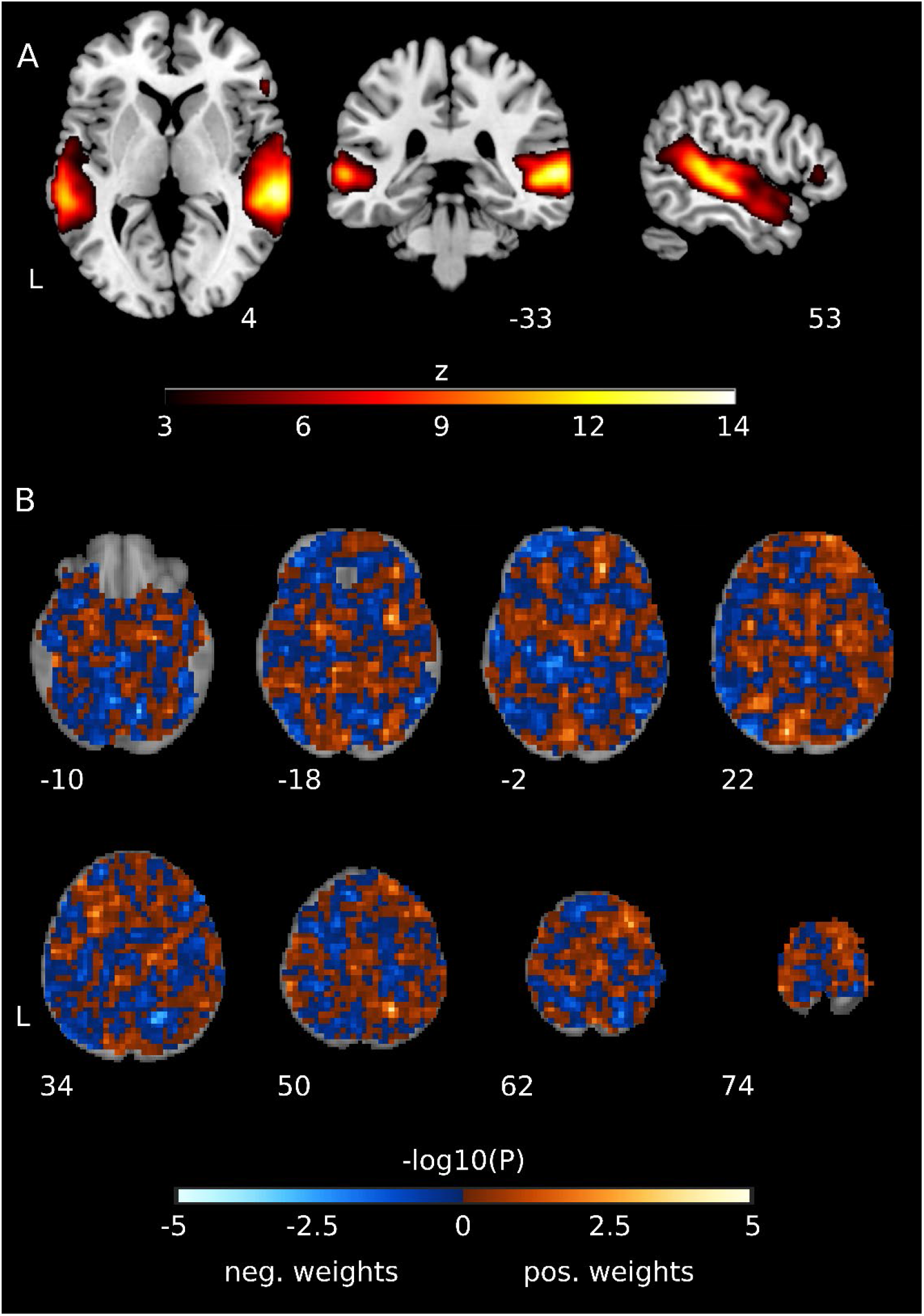
A. A network centered on the bilateral superior temporal gyrus which provided the best performance during the multivariate classification of responders and non-responders. B. p-values of the individual voxel weights of the SVM estimated using the margin-aware statistic and analytical approximation of the null-distribution [41]. P-values are shown unthresholded as the analysis is multivariate and therefore all voxels – and not only the most significant ones – always contribute to the classification task.

**Figure 3:**
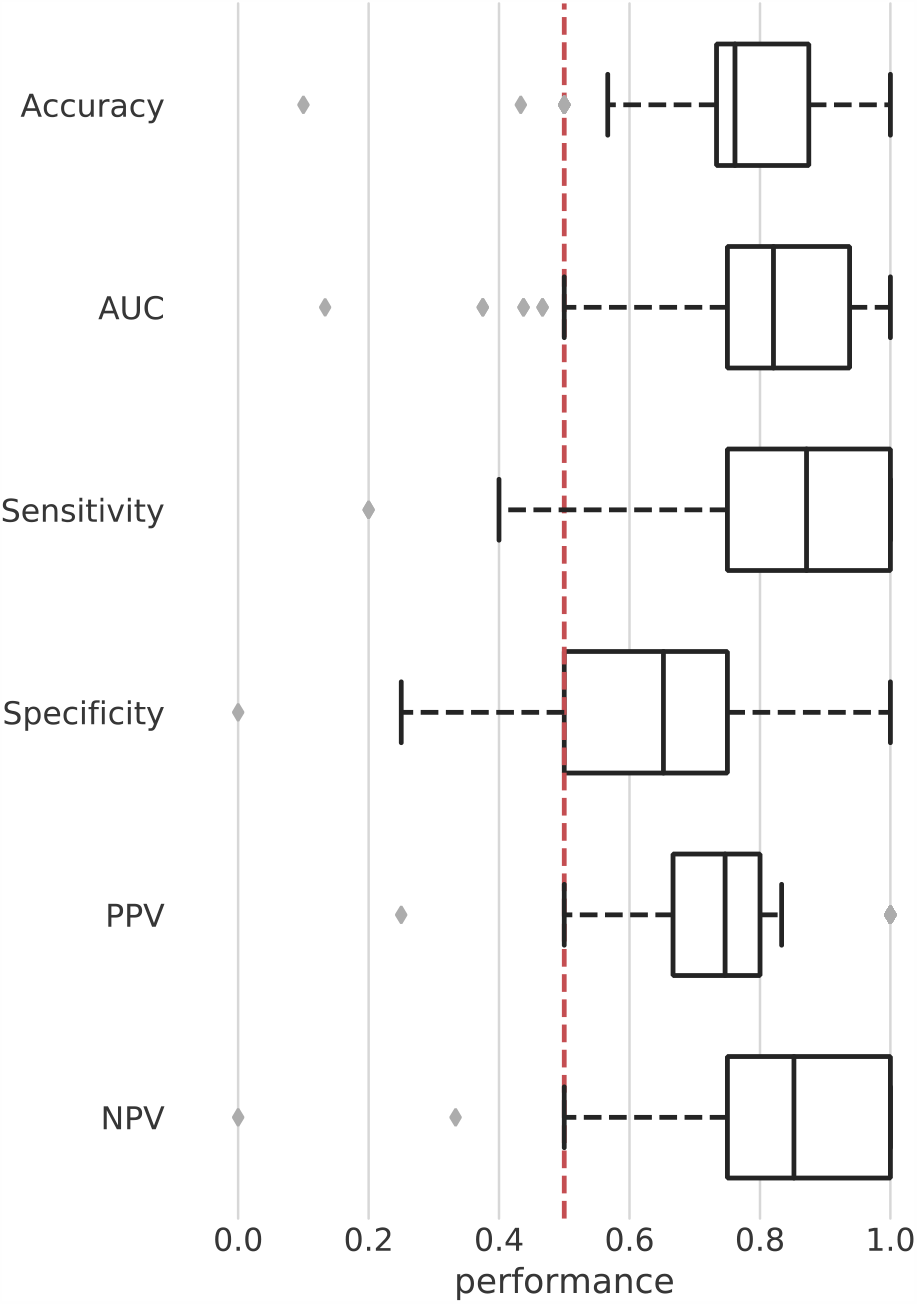
Cross-validated performance estimates of the best performing network during classification (Figure 2). Boxplots show the mean and interquartile range (IQR) of the individual performance distributions. The mean instead of the median is shown because it was also used and reported as final performance of the network. Red dotted line indicates approximate chance-level. However, statistically, deviation from chance-level and FWE-correction were estimated through synchronized permutations.

## Discussion

In this study we investigated the possibility of using pre-treatment rs-fMRI data as a biomarker to predict trauma-focused psychotherapy response in youth with (partial) PTSD. We examined prediction both on the group- and individual-level. In our study, a network centered on the bilateral STG could distinguish between responders and non-responders on the individual-level, with an accuracy of 76.2%. We further found increased connectivity between the left FPN and a sensorimotor network in non-responders on the group-level. To our knowledge this is the first study to examine the prediction of treatment-response using rs-fMRI data in youth with PTSD. Together our results provide a first proof-of-concept for the utility of rs-fMRI as a biomarker for treatment-response in youth with PTSD.

Our findings indicate increased pre-treatment connectivity between the left FPN and sensorimotor network in trauma-focused psychotherapy non-responders. The FPN is highly integrated with other brain networks and has a comprehensive role in attention, working memory and decision making by flexibly interacting with other brain networks [10]. Abnormal recruitment of other brain networks into the FPN is linked with deficits in these cognitive processes and has been associated with multiple psychiatric disorders [10]. More specifically, increased connectivity between the FPN and a sensorimotor network has been found in youth with autism (ASD) and attention-deficit/hyperactivity disorder (ADHD) [36]. While speculative at this point, abnormal recruitment of the sensorimotor network into the FPN in non-responders might be related to deficient cognitive processes resulting in suboptimal engagement in trauma-focused psychotherapy and poor treatment response. To test this hypothesis, future research could address functional connectivity patterns of the FPN together with neurocognitive tests before and after treatment and use repeated transcranial magnetic stimulation to directly influence FPN connectivity [23]. Such an approach could eventually delineate clinical relevance and might identify promising targets for non-invasive stimulation-based interventions [42].

The ICN yielding significant classification performance was centered on the STG. A growing number of studies have shown structural and functional abnormalities in the STG in PTSD patients [43-45]. Based on electrical stimulation of the area, Engdahl and colleagues [44] have suggested that STG abnormalities may be associated with re-experiencing symptoms. Others have suggested a relationship between STG abnormalities and dissociative symptoms in PTSD patients [43]. Interestingly, we have previously shown a positive correlation between STG activation and trauma-focused psychotherapy response in adults with PTSD [45].

Previous studies utilizing ML methods, however, did not identify network connectivity of the STG as an accurate predictor of treatment response. In adults treated with prolonged exposure, Etkin, et al. [23] found a classification accuracy of >85%, using a combination of pre-treatment rs-fMRI connectivity within the ventral attention network and delayed recall performance in a verbal memory task. In another study, pre-treatment functional connectivity within- and between-the default mode, dorsal attention, cingulo-opercular, salience, and central executive network during task-free fMRI predicted response to TF-CBT with an accuracy of 71.4% [20]. Finally, we have previously shown the feasibility of the same approach as outlined here to predict response to trauma-focused therapy in veterans with PTSD with 81.4% accuracy [17], with an ICN centered on the pre-supplementary motor area providing the best predictive accuracy.

At present, it remains unclear why our findings on classification accuracy differ from findings in adult PTSD. Studies in adults have reported different networks/functional connectivity estimates than identified here and have found classification accuracies which mostly exceeded accuracy found in the current study. One possibility is that, with inclusion of both PTSD and partial PTSD patients, clinical heterogeneity increased, resulting in lower classification accuracy. Another possibility is that neurodevelopmental trajectories add to heterogeneity and might reduce classification accuracy, as previous studies in youth with PTSD using rs-fMRI have shown neurodevelopmental effects on network connectivity and we included youth with a relatively wide age range. These hypotheses require further investigation, including longitudinal studies of youth with PTSD which develop into adulthood. While the current individual-level classification findings differ from adults, it is reassuring that the application of the same approach to treatment-response classification as reported here has been associated with significant classification accuracies in adults multiple times, even for a different disorder [17,46].

There is a difference between the findings observed on the group- and on the individual-level. While there was no difference in within-network connectivity for any ICN between responders and non-responders on the group-level, there was a network significantly predictive on the individual-level. The opposite was true for the between-network connectivity. These discrepancies can be explained by the fact that a significant p-value in group-comparisons does not have to imply the ability to distinguish between patients on the individual-level because of low effect sizes of the difference [22]. In addition, both analyses have different goals and therefore can identify different ICNs: group-level analyses focus on determining localized average differences between groups while individual-level analyses utilize all multivariate data to determine a model which provides the highest prediction [47]. This clearly marks the importance of performing individual-level prediction studies as these may improve clinical decision making in the future and may lead to independent results from group-level studies.

Although classification accuracy exceeded chance-level performance, it still falls below the APA proposed threshold for clinical applicability of biomarkers [48]. The suggested combination of >80% sensitivity, specificity, and PPV is useful as guidance for research, but clinical utility should preferably be based on a cost-benefit analysis [49]. As the current clinical standard is to offer trauma-focused psychotherapy to all youth with PTSD, a biomarker which reliably identifies treatment non-responders could aid clinical decision making. This would correspond to a classifier with high specificity, but also with sensitivity of at least 80% to prevent classifying all patients as non-responders. If a-priory chances of treatment non-response are high, clinicians together with patients and their caregivers, could decide to abstain from initiating trauma-focused psychotherapy and search for alternative treatments with higher chances of success. This may help to prevent the unnecessary burden of failed treatment trials. This study shows the promise of combining machine learning and rs-fMRI to identify prospective biomarkers for treatment response in youth with PTSD but the observed performance makes a clinical application difficult at the current state.

Several limitations of this study should be noted. First, the sample size in the current study is low. This has an impact on the certainty of the estimated performance of the individual-level analysis. Cross-validation can lead to high variance in performance estimates if applied to studies with low sample sizes [50]. To increase the confidence in the presented results, we followed best-practices for the field [35] and utilized a permutation test corrected for multiple comparisons which can provide a valid statistical control for the performance measures [50]. However, only with larger sample sizes can these problems be fully addressed and therefore the current study can only be regarded as a first step for further individual-level prediction studies in youth with PTSD. Larger sample sizes at the same time may increase clinical heterogeneity and limit classification performance as well [22]. Second, although the majority (82.5%) of included youth had a full PTSD diagnosis, the remaining 17.5% had a partial PTSD diagnosis. Including youth with partial PTSD increased clinical heterogeneity. Increased clinical heterogeneity might have lowered overall treatment response due to a floor effect and might have lowered prediction accuracy. However, by including youth with partial PTSD, our sample better reflects the real-life clinical setting, which adds to the ecological validity of our findings. Third, youth were randomized to receive either TF-CBT or EMDR, and both treatment conditions were collapsed for the current analysis. Due to limited power it was not feasible to examine differences between treatment responders and non-responders separately for both treatments or examine specific predictors for each treatment separately. Importantly, efficacy of both treatments has been shown comparable in an RCT with considerable sample overlap with the current study [6]. Finally, our study had substantial drop-out, as 18% of randomized patients were lost to follow-up. Although such dropout rates reflect routine clinical practice and treatment completers and non-completers did not differ on baseline characteristics, there is a possibility that drop-out could have influenced our findings through attrition bias.

The present study demonstrates that increased resting-state connectivity between the FPN and a sensorimotor network can distinguish trauma-focused psychotherapy responders from non-responders on the group-level. Future studies could examine if these network patterns are potential targets for (non-invasive) neuromodulation interventions to reduce PTSD symptoms in afflicted youth. We further show that resting-state connectivity patterns in a network centered on the bilateral STG are capable of predicting trauma-focused psychotherapy response in youth with PTSD. These proof-of-concept findings emphasize the feasibility of combining machine learning analysis and rs-fMRI to identify prospective biomarkers for treatment response. However, before translation to clinical practice can commence, future research should aim to test the robustness and generalizability of these findings in larger independent cohorts.

## Data Availability

Data available upon reasonable request.

## References

1 Alisic E, Zalta AK, van Wesel F, Larsen SE, Hafstad GS, Hassanpour K, et al. Rates of post-traumatic stress disorder in trauma-exposed children and adolescents: meta-analysis. The British journal of psychiatry : the journal of mental science. 2014;204:335–40.

2 American Psychiatric Association. Diagnostic and statistical manual of mental disorders (DSM-5®). American Psychiatric Pub; 2013.

3 Carrion VG, Weems CF, Ray R, Reiss AL. Toward an empirical definition of pediatric PTSD: the phenomenology of PTSD symptoms in youth. Journal of the American Academy of Child and Adolescent Psychiatry. 2002;41(2):166–73.

4 Molnar BE, Berkman LF, Buka SL. Psychopathology, childhood sexual abuse and other childhood adversities: relative links to subsequent suicidal behaviour in the US. Psychological medicine. 2001;31(6):965–77.

5 Morina N, Koerssen R, Pollet TV. Interventions for children and adolescents with posttraumatic stress disorder: A meta-analysis of comparative outcome studies. Clinical Psychology Review. 2016;47:41–54.

6 Diehle J, Opmeer BC, Boer F, Mannarino AP, Lindauer RJ. Trauma-focused cognitive behavioral therapy or eye movement desensitization and reprocessing: what works in children with posttraumatic stress symptoms? A randomized controlled trial. European child & adolescent psychiatry. 2015;24(2):227–36.

7 Goldbeck L, Muche R, Sachser C, Tutus D, Rosner R. Effectiveness of trauma-focused cognitive behavioral therapy for children and adolescents: A randomized controlled trial in eight German mental health clinics. Psychotherapy and Psychosomatics. 2016;85(3):159–70.

8 Akiki TJ, Averill CL, Abdallah CG. A network-based neurobiological model of PTSD: evidence from structural and functional neuroimaging studies. Current Psychiatry Reports. 2017;19(11):81.

9 Sripada RK, King AP, Garfinkel SN, Wang X, Sripada CS, Welsh RC, et al. Altered resting-state amygdala functional connectivity in men with posttraumatic stress disorder. Journal of psychiatry & neuroscience: JPN. 2012;37(4):241.

10 Menon V. Large-scale brain networks and psychopathology: a unifying triple network model. Trends in cognitive sciences. 2011;15(10):483–506.

11 Bluhm RL, Williamson PC, Osuch EA, Frewen PA, Stevens TK, Boksman K, et al. Alterations in default network connectivity in posttraumatic stress disorder related to early-life trauma. Journal of psychiatry & neuroscience: JPN. 2009;34(3):187.

12 Liu Y, Li L, Li B, Feng N, Li L, Zhang X, et al. Decreased triple network connectivity in patients with recent onset post-traumatic stress disorder after a single prolonged trauma exposure. Scientific reports. 2017;7(1):1–10.

13 Weems CF, Russell JD, Neill EL, McCurdy BH. Annual research review: Pediatric posttraumatic stress disorder from a neurodevelopmental network perspective. Journal of Child Psychology and Psychiatry. 2019;60(4):395–408.

14 Patriat R, Birn RM, Keding TJ, Herringa RJ. Default-mode network abnormalities in pediatric posttraumatic stress disorder. Journal of the American Academy of Child & Adolescent Psychiatry. 2016;55(4):319–27.

15 Menon V. Developmental pathways to functional brain networks: emerging principles. Trends in cognitive sciences. 2013;17(12):627–40.

16 Duval ER, Sheynin J, King AP, Phan KL, Simon NM, Martis B, et al. Neural function during emotion processing and modulation associated with treatment response in a randomized clinical trial for posttraumatic stress disorder. Depression and Anxiety. 2020.

17 Zhutovsky P, Thomas RM, Olff M, van Rooij SJH, Kennis M, van Wingen GA, et al. Individual prediction of psychotherapy outcome in posttraumatic stress disorder using neuroimaging data. Transl Psychiatry. 2019;9(1):326.

18 Zantvoord JB, Diehle J, Lindauer RJ. Using neurobiological measures to predict and assess treatment outcome of psychotherapy in posttraumatic stress disorder: systematic review. Psychotherapy and psychosomatics. 2013;82(3):142–51.

19 Fonzo GA, Goodkind MS, Oathes DJ, Zaiko YV, Harvey M, Peng KK, et al. PTSD psychotherapy outcome predicted by brain activation during emotional reactivity and regulation. American Journal of Psychiatry. 2017;174(12):1163–74.

20 Korgaonkar MS, Chakouch C, Breukelaar IA, Erlinger M, Felmingham KL, Forbes D, et al. Intrinsic connectomes underlying response to trauma-focused psychotherapy in post-traumatic stress disorder. Transl Psychiatry. 2020;10(1):270.

21 Cisler JM, Sigel BA, Kramer TL, Smitherman S, Vanderzee K, Pemberton J, et al. Modes of largescale brain network organization during threat processing and posttraumatic stress disorder symptom reduction during TF-CBT among adolescent girls. Plos one. 2016;11(8):e0159620.

22 Arbabshirani MR, Plis S, Sui J, Calhoun VD. Single subject prediction of brain disorders in neuroimaging: Promises and pitfalls. Neuroimage. 2017;145(Pt B):137–65.

23 Etkin A, Maron-Katz A, Wu W, Fonzo GA, Huemer J, Vertes PE, et al. Using fMRI connectivity to define a treatment-resistant form of post-traumatic stress disorder. Sci Transl Med. 2019;11(486).

24 Nader K, Kriegler J, Blake D, Pynoos R, Newman E, Weather F. Clinician administered PTSD scale, child and adolescent version. White River Junction, VT: National Center for PTSD. 1996;156.

25 Verlinden E, van Laar YL, van Meijel EP, Opmeer BC, Beer R, de Roos C, et al. A parental tool to screen for posttraumatic stress in children: first psychometric results. J Trauma Stress. 2014;27(4):492–5.

26 Stein MB, Walker JR, Hazen AL, Forde DR. Full and partial posttraumatic stress disorder: findings from a community survey. Am J Psychiatry. 1997;154(8):1114–9.

27 Zantvoord JB, Ensink JB, op den Kelder R, Wessel AM, Lok A, Lindauer RJ. Pretreatment cortisol predicts trauma-focused psychotherapy response in youth with (partial) posttraumatic stress disorder. Psychoneuroendocrinology. 2019;109:104380.

28 Zantvoord JB, Zhutovsky P, Ensink JBM, Op den Kelder R, van Wingen GA, Lindauer RJL. Trauma-focused psychotherapy response in youth with posttraumatic stress disorder is associated with changes in insula volume. J Psychiatr Res. 2021;132:207–14.

29 Chorpita BF, Yim L, Moffitt C, Umemoto LA, Francis SE. Assessment of symptoms of DSM-IV anxiety and depression in children: a revised child anxiety and depression scale. Behav Res Ther. 2000;38(8):835–55.

30 Power JD, Mitra A, Laumann TO, Snyder AZ, Schlaggar BL, Petersen SE. Methods to detect, characterize, and remove motion artifact in resting state fMRI. Neuroimage. 2014;84:320–41.

31 Pruim RHR, Mennes M, van Rooij D, Llera A, Buitelaar JK, Beckmann CF. ICA-AROMA: A robust ICA-based strategy for removing motion artifacts from fMRI data. Neuroimage. 2015;112:267–77.

32 Lindquist MA, Geuter S, Wager TD, Caffo BS. Modular preprocessing pipelines can reintroduce artifacts into fMRI data. Hum Brain Mapp. 2019;40(8):2358–76.

33 Biswal BB, Mennes M, Zuo XN, Gohel S, Kelly C, Smith SM, et al. Toward discovery science of human brain function. Proc Natl Acad Sci U S A. 2010;107(10):4734–9.

34 Beckmann CF, Smith SM. Probabilistic independent component analysis for functional magnetic resonance imaging. IEEE Trans Med Imaging. 2004;23(2):137–52.

35 Poldrack RA, Huckins G, Varoquaux G. Establishment of Best Practices for Evidence for Prediction: A Review. JAMA Psychiatry. 2019.

36 Cerliani L, Mennes M, Thomas RM, Di Martino A, Thioux M, Keysers C. Increased Functional Connectivity Between Subcortical and Cortical Resting-State Networks in Autism Spectrum Disorder. JAMA Psychiatry. 2015;72(8):767–77.

37 Du Y, Fan Y. Group information guided ICA for fMRI data analysis. Neuroimage. 2013;69:157–97.

38 Salman MS, D.Y, Lin D, Fu Z, Fedorov A, Damaraju E, et al. Group ICA for identifying biomarkers in schizophrenia: ‘Adaptive’ networks via spatially constrained ICA show more sensitivity to group differences than spatio-temporal regression. Neuroimage Clin. 2019;22:101747.

39 Smith SM, Nichols TE. Threshold-free cluster enhancement: addressing problems of smoothing, threshold dependence and localisation in cluster inference. Neuroimage. 2009;44(1):83–98.

40 Cortes C, Vapnik V. Support-vector networks. Machine Learning. 1995;20(3):273–97.

41 Gaonkar B, Shinohara R, Davatzikos C, Alzheimers Disease Neuroimaging I. Interpreting support vector machine models for multivariate group wise analysis in neuroimaging. Med Image Anal. 2015;24(1):190–204.

42 Fonzo GA, Goodkind MS, Oathes DJ, Zaiko YV, Harvey M, Peng KK, et al. Selective Effects of Psychotherapy on Frontopolar Cortical Function in PTSD. Am J Psychiatry. 2017;174(12):1175–84.

43 Lanius RA, Williamson PC, Boksman K, Densmore M, Gupta M, Neufeld RW, et al. Brain activation during script-driven imagery induced dissociative responses in PTSD: a functional magnetic resonance imaging investigation. Biol Psychiatry. 2002;52(4):305–11.

44 Engdahl B, Leuthold AC, Tan HR, Lewis SM, Winskowski AM, Dikel TN, et al. Post-traumatic stress disorder: a right temporal lobe syndrome? J Neural Eng. 2010;7(6):066005.

45 Lindauer RJ, Booij J, Habraken JB, van Meijel EP, Uylings HB, Olff M, et al. Effects of psychotherapy on regional cerebral blood flow during trauma imagery in patients with post-traumatic stress disorder: a randomized clinical trial. Psychological medicine. 2008;38(4):543–54.

46 van Waarde JA, Scholte HS, van Oudheusden LJ, Verwey B, Denys D, van Wingen GA. A functional MRI marker may predict the outcome of electroconvulsive therapy in severe and treatment-resistant depression. Mol Psychiatry. 2015;20(5):609–14.

47 Bzdok D, Ioannidis JPA. Exploration, Inference, and Prediction in Neuroscience and Biomedicine. Trends Neurosci. 2019;42(4):251–62.

48 First MB, Drevets WC, Carter C, Dickstein DP, Kasoff L, Kim KL, et al. Clinical Applications of Neuroimaging in Psychiatric Disorders. Am J Psychiatry. 2018;175(9):915–16.

49 Pepe MS, Janes H, Li CI, Bossuyt PM, Feng Z, Hilden J. Early-Phase Studies of Biomarkers: What Target Sensitivity and Specificity Values Might Confer Clinical Utility? Clin Chem. 2016;62(5):737–42.

50 Varoquaux G. Cross-validation failure: Small sample sizes lead to large error bars. Neuroimage. 2018;180(Pt A):68–77.

