## Supplementary Materials for "Individual prediction of trauma-focused psychotherapy response in youth with posttraumatic stress disorder using resting-state functional connectivity"

**Trauma-exposed controls**

To prevent overfitting in the main analyses, we utilized an independent sample of trauma-exposed controls (TEC) to identify our intrinsic connectivity network (ICN) templates . TEC were aged between 8 and 18 years and were able to understand the Dutch language. TEC were recruited between June 2011 and September 2018 through local elementary- and high schools by researcher JBZ, RodK and JBME. Exposure to traumatic events were validated according to A1 and A2 criteria of DSM-IV-TR [2] using the life-events checklist of the Clinician-Administered PTSD Scale for Children and Adolescents (CAPS-CA) semi-structured interview [3]. TEC were excluded if they met PTSD or partial PTSD diagnosis using both the CAPS-CA and caregiver reports from the PTSD scale of the Anxiety Disorders Interview Schedule – Parent Version (ADIS-P) [4] or had a CAPS-CA total score of >20 points. Additional exclusion criteria were: acute suicidality, IQ<70, pregnancy, neurological disorders or serious medical illnesses or meeting the criteria of one of the following diagnosis: psychotic disorders, substance-use disorder or pervasive developmental disorder. 21 TEC were scanned on the same scanner using the same parameters and protocol as the (partial)-PTSD patients described in the main manuscript.

The resting-state functional magnetic resonance imaging (rs-fMRI) data of the TEC was preprocessed according to the exact same procedures as described in the main manuscript. Application of the same quality control procedures led to the exclusion of two TEC due to registration failures and two TEC due to excess motion, leading to a final included sample of 17 TEC as reported in the main manuscript. The included TEC did not differ from the included PTSD patients in age (M: 14, SD: 3.57, $t(55) = 1.53$, p = 0.133), gender (64.7% female, $X^2(1) = 0.0005$, p = 0.983), age at trauma (M; 12.13, SD 3.52, $t(52) = 1.87$, p = 0.063), time since trauma (M; 2.07,

SD 1.98, $t(52)$ =1.039, p = 0.304), and motion as estimated via mean framewise displacement[5] (M: 0.15, SD: 0.04, $t(55)$ = -1.77, p = 0.08) but did differ in type of trauma exposure $X^2(4)$ = 15.998, p = 0.003 with relatively more accidents and other trauma and less sexual abuse and domestic/community violence in the TEC group.

**Functional data preprocessing**

For each participant, a reference rs-fMRI volume and its skull-stripped version were generated using custom methodology of fMRIPrep. This reference image was then co-registered to the corresponding MRI scan using boundary-based registration [6]. Head-motion correction with respect to the reference was estimated before any spatiotemporal filtering using MCFLIRT [7]. The rs-fMRI scans were normalized to MNI space combining all spatial transformations (head-motion correction, co-registration and normalization) into one single step using Lanczos interpolation [8].

**Identification of intrinsic connectivity networks**

The meta-ICA procedure was implemented as follows: we repeatedly (n=25) and randomly selected 15 out of our 17 TEC and computed a temporally-concatenated ICA to identify spatially independent components (70 components each, i.e. 25 * 70 = 1750 spatial components in total). Then, we concatenated all spatial components across all individual ICA runs and calculated a final meta-ICA (70 components) leading to a set of robust spatial components to consider for identification of ICNs.

To identify valid ICNs, we employed a semi-automatic approach [9]. In a first stage, we assessed all spatial components visually, focusing on overlap with GM and overlap with previously identified ICNs. This led to the exclusion of 20 components. In a second stage, we computed the average spatial correlation between each of the meta-ICA components and the maximally correlated spatial components of each individual ICA run [9]. Such a measure represents the reproducibility of the meta-ICA components across all individual ICA runs. We excluded all components with a correlation <0.6, leading to the exclusion of 2 additional components.

**Individual-level analyses**

To assess statistically whether the averaged, cross-validated balanced accuracies allowed for better-than-chance performance and to correct for the number of classifications performed, we utilized synchronized permutation tests of the maximum statistic. To this end we randomly permuted the classification labels associated with our data (same permutations for each ICN and the between-ICN connectivity, n=2000) [10], estimated the maximum performance for each of the permutations across all the included ICNs/between-ICN connectivity measures and used this estimated null-distribution of the maximum statistic to correct for familywise-error of the p-values of the individual performances of each of our classifications.

We also assessed which features (voxel values of individual ICNs or individual between-ICN connectivity) were important for the SVM classification by calculating p-values for each weight of the classifier. The p-values were estimated using an analytical approximation of a permutation procedure from a combination of the weights and the size of the margin of the SVM [11]. The p-values were computed after the classifier was applied to the entire data set (no cross-validation) and are intended for post-hoc visualization purposes only.

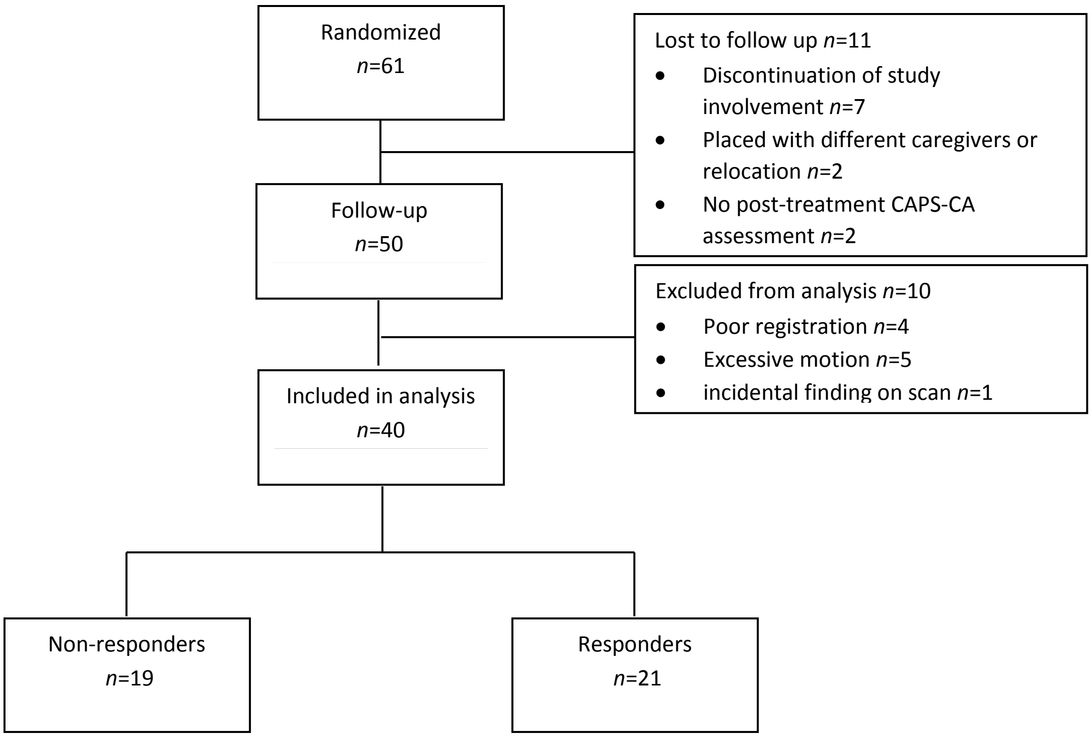


**Figure S1**: Flow diagram of included patients. Response is defined as ≥30% reduction in CAPS-CA total score from pre- to post-treatment.

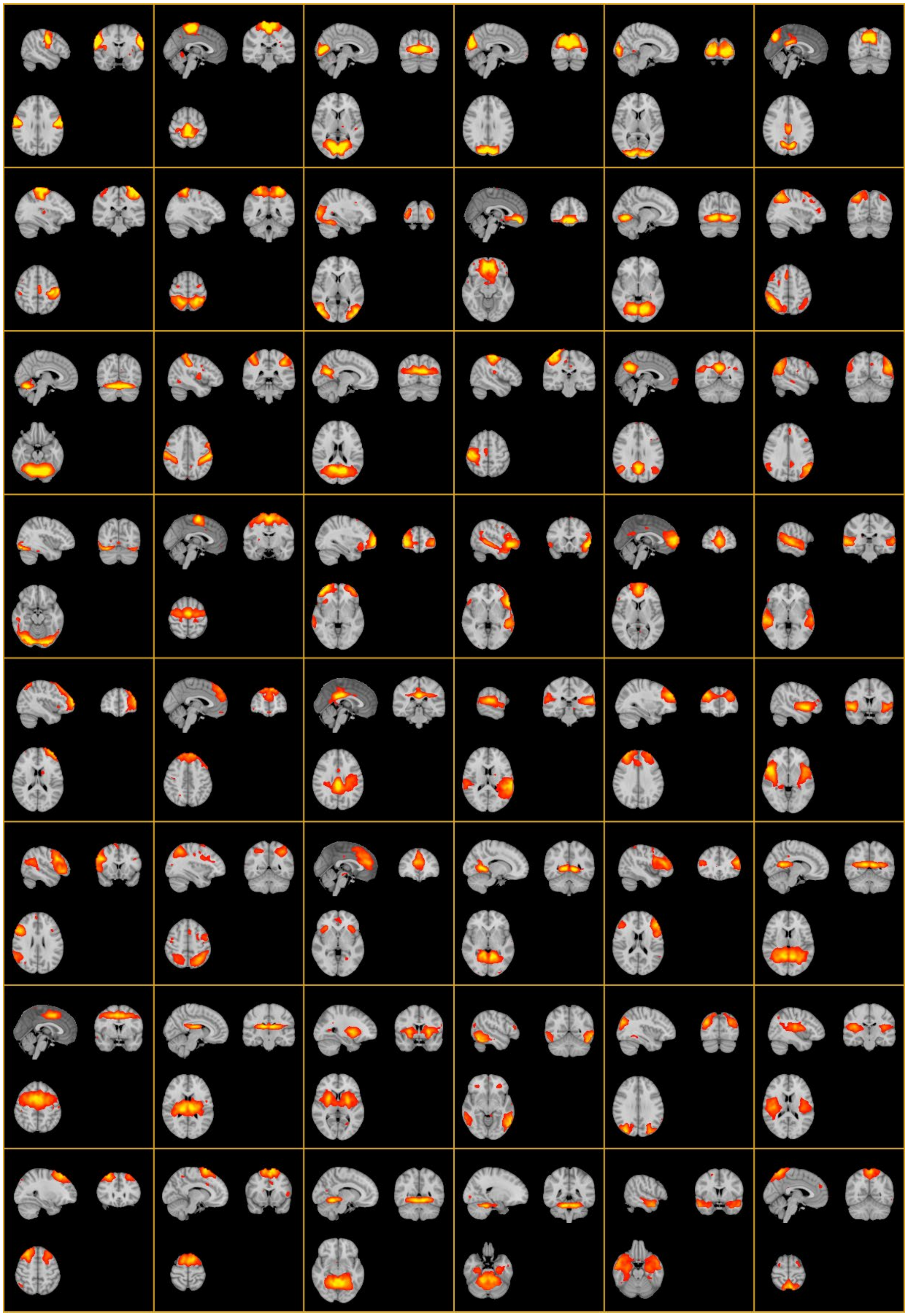

**Figure S2:** All 48 included intrinsic connectivity networks (ICNs) estimated through the application of meta-independent component analyses (ICA) with 70 (group-)components. ICNs were identified utilizing a semi-automatic approach consisting of visual assessment and calculation of average spatial correlation coefficients between each of the meta-ICA spatial components and their corresponding maximally correlated individual-ICA components. Components with an average correlation <0.6 were removed. The right hemisphere is plotted on the left.

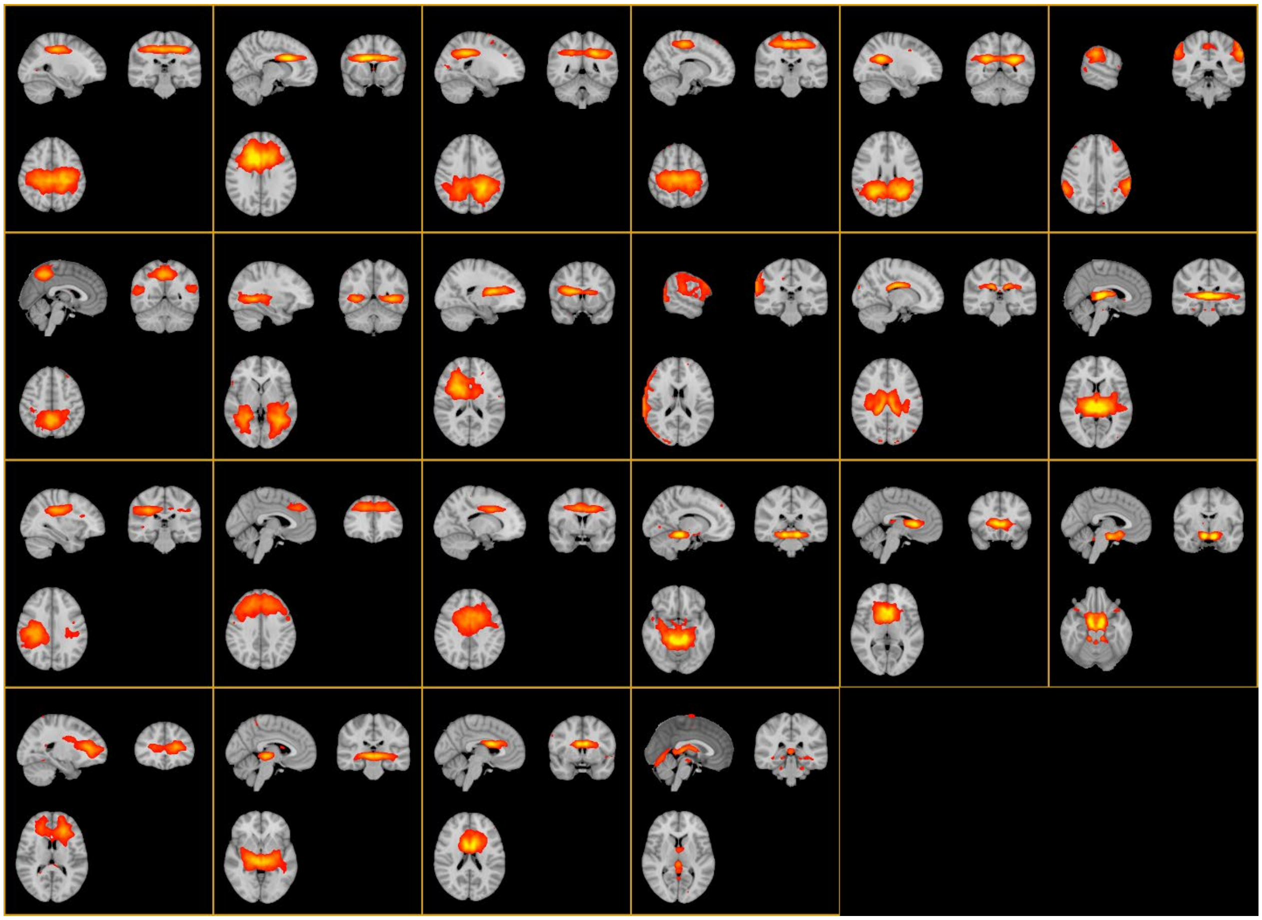

**Figure S3:** All 22 excluded components. Either excluded because of their overlap with white matter/cerebrospinal fluid or because of their low average spatial correlation (<0.6) between the meta-ICA group-component and its maximally correlated individual-ICA components. The right hemisphere is plotted on the left.

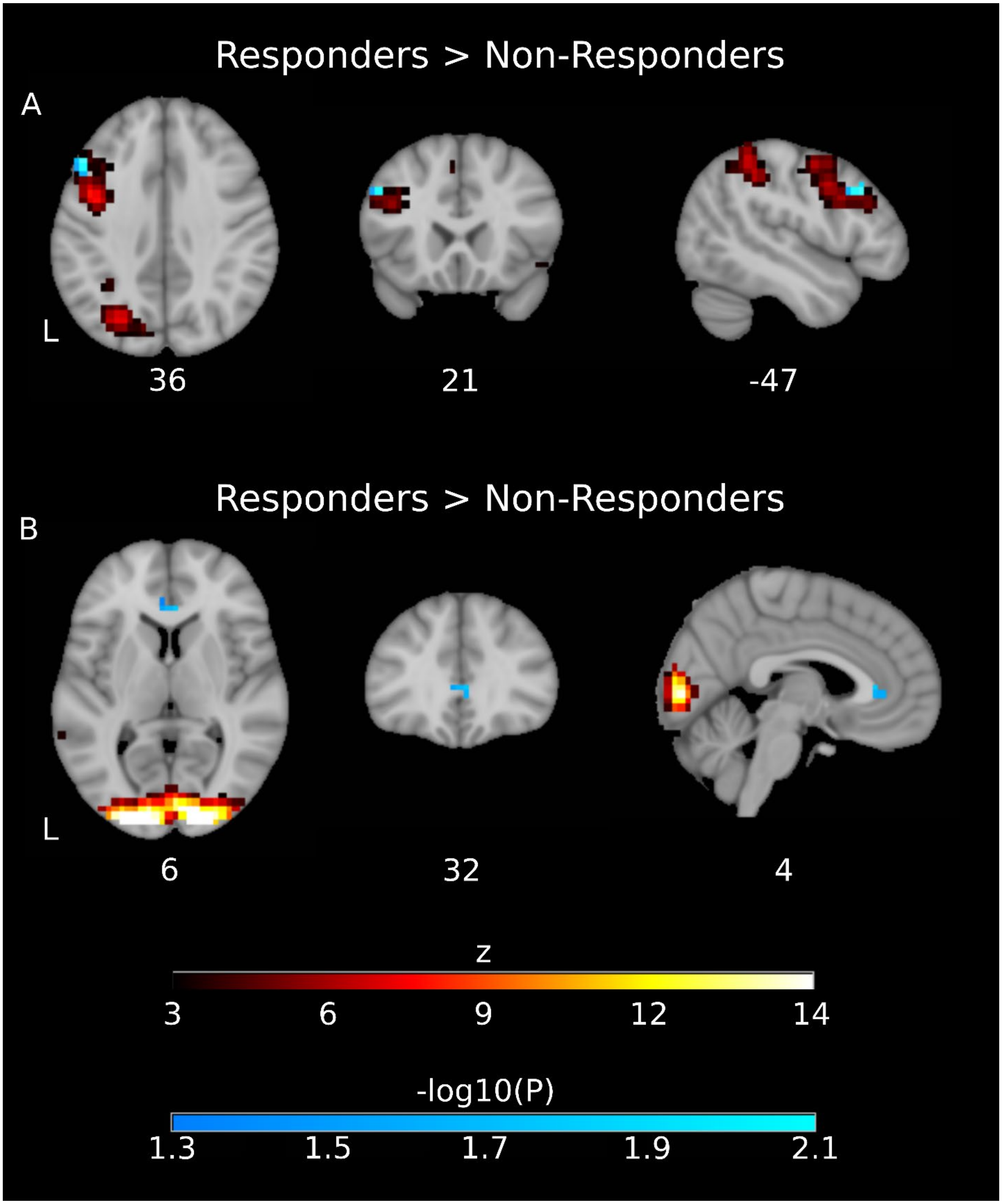


**Figure S4:** Exploratory group-level analyses comparing responders to non-responders for within-ICN connectivity without correction for multiple-comparisons for the number of investigated components. **A.** increased within-ICN connectivity in the left frontoparietal ICN (same as in Figure

1 of the main manuscript) for responders over non-responders. **B.** increased within-ICN connectivity in the visual (occipital) ICN for responders over non-responders.

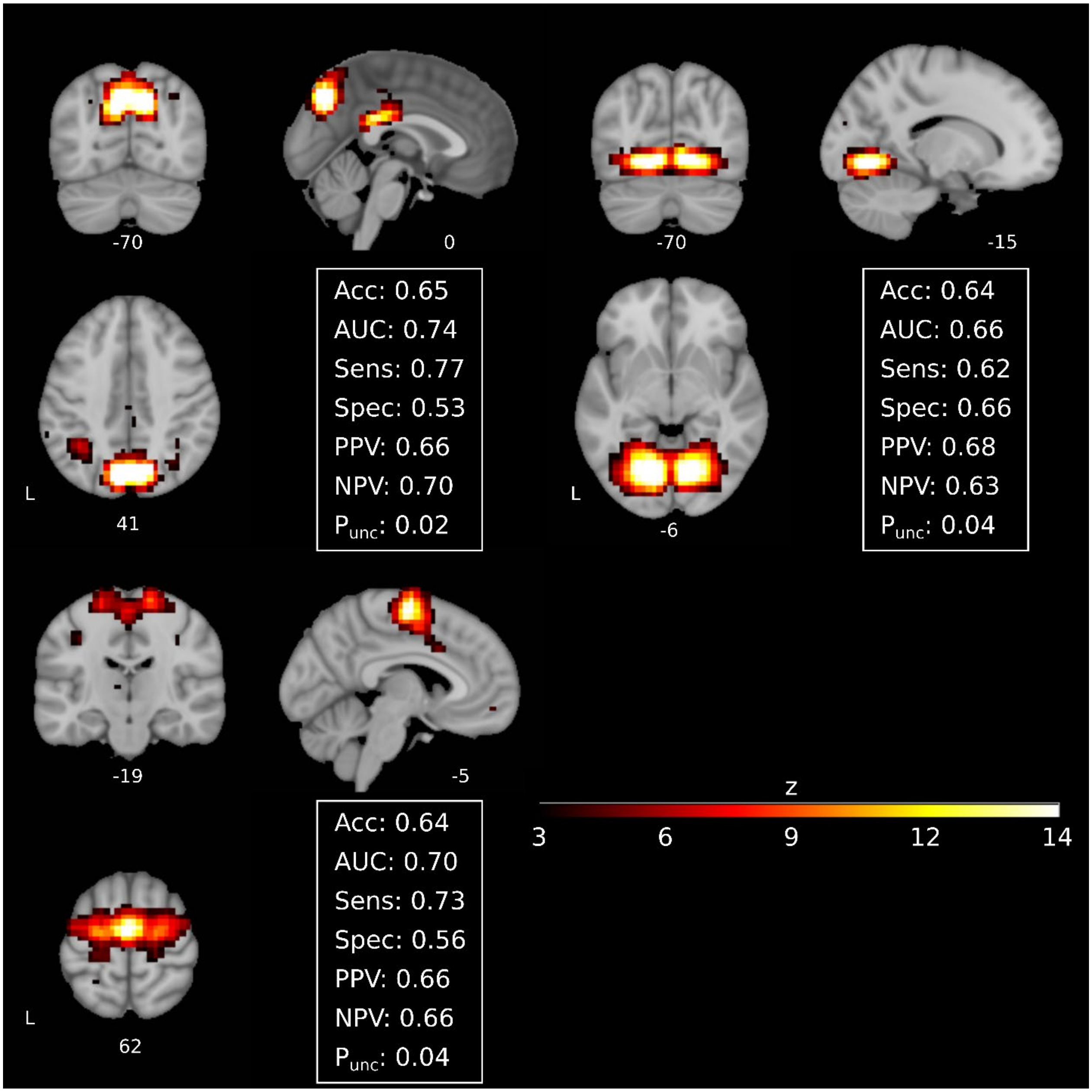


**Figure S5:** Exploratory classification analyses without additional correction for the number of investigated components. Shows performance of three additional networks which show better-than-chance performance in this exploratory investigation.

### References


1 Poldrack RA, Huckins G, Varoquaux G. Establishment of Best Practices for Evidence for Prediction: A Review. JAMA Psychiatry. 2019.

2 American Psychiatric Association. Diagnostic and statistical manual of mental disorders. (4th ed., Text Revision) ed. American Psychiatric Association: Washington, DC; 2000.

3 Nader K, Kriegler J, Blake D, Pynoos R, Newman E, Weather F. Clinician administered PTSD scale, child and adolescent version. White River Junction, VT: National Center for PTSD. 1996;156.

4 Verlinden E, van Meijel EP, Opmeer BC, Beer R, de Roos C, Bicanic IA, et al. Characteristics of the Children's Revised Impact of Event Scale in a clinically referred Dutch sample. Journal of Traumatic Stress. 2014;27(3):338-44.

5 Power JD, Mitra A, Laumann TO, Snyder AZ, Schlaggar BL, Petersen SE. Methods to detect, characterize, and remove motion artifact in resting state fMRI. Neuroimage. 2014;84:320-41.

6 Greve DN, Fischl B. Accurate and robust brain image alignment using boundary-based registration. Neuroimage. 2009;48(1):63-72.

7 Jenkinson M, Bannister P, Brady M, Smith S. Improved optimization for the robust and accurate linear registration and motion correction of brain images. Neuroimage. 2002;17(2):825-41.

8 Lanczos C. Evaluation of Noisy Data. Journal of the Society for Industrial and Applied Mathematics: Series B, Numerical Analysis. 1964;1:76-85.

9 Cerliani L, Mennes M, Thomas RM, Di Martino A, Thioux M, Keysers C. Increased Functional Connectivity Between Subcortical and Cortical Resting-State Networks in Autism Spectrum Disorder. JAMA Psychiatry. 2015;72(8):767-77.

10 Ojala M, Garriga GC. Permutation Tests for Studying Classifier Performance. J Mach Learn Res. 2010;11:1833-63.

11 Gaonkar B, Shinohara R, Davatzikos C, Alzheimers Disease Neuroimaging I. Interpreting support vector machine models for multivariate group wise analysis in neuroimaging. Med Image Anal. 2015;24(1):190-204.
